# Using the COVID-19 to influenza ratio to estimate the numbers of symptomatic COVID-19 cases in Wuhan prior to the lockdown

**DOI:** 10.1101/2020.04.26.20075937

**Authors:** Zhanwei Du, Benjamin J. Cowling, Lauren Ancel Meyers

**Affiliations:** The University of Texas at Austin, Austin, Texas 78712, The United States of America; The University of Hong Kong, 7 Sassoon Rd, Hong Kong SAR, China; Santa Fe Institute, Santa Fe, New Mexico, The United States of America

## Abstract

A recent study tested 45 throat swabs taken from adults over age 30 who sought outpatient care at one of two central Wuhan hospitals for influenza-like-illness between December 30, 2019 and January 19, 2020. Although none were confirmed COVID-19 cases, nine retrospectively tested positive for the virus. Using the fact that Wuhan has 393 other hospitals, we extrapolate the total number of undetected cases of symptomatic COVID-19 in adults during this period. we estimate that there were 5,558 [95% CI: 2,761-9,864] adults with symptomatic COVID-19 infections in Wuhan between December 30th and January 19th, 2020.

---

Only 422 cases of COVID-19 were reported in mainland China before January 22, 2020 (1). A recent study (2) tested 45 throat swabs taken from adults over age 30 who sought outpatient care at one of two central Wuhan hospitals for influenza-like-illness between December 30, 2019 and January 19, 2020. Although none were confirmed COVID-19 cases, nine retrospectively tested positive for the virus. Using the fact that Wuhan has 393 other hospitals, we extrapolate the total number of undetected cases of symptomatic COVID-19 in adults during this period.

In addition to the nine COVID-19 positive samples, seven others tested positive for influenza (2). Coupling the ratio between COVID-19 and influenza positive tests with an estimate for influenza prevalence in Wuhan during the same time derived from surveillance data (see Appendix), we estimate that there were 5,558 [95% CI: 2,761-9,864] adults with symptomatic COVID-19 infections in Wuhan between December 30th and January 19th, ranging from 75 cases [95% CI: 37-133] in suburban Hannan to 711 cases [95% CI: 353-1,262] in central Wuchang (Figure). Estimates for the epidemic doubling time for COVID-19 in Hubei Province have ranged from 5.2 (3) to 7.31 days (4). These two values suggest a total of 10,236 (95% CI: 3,797-25,864) or 9,934 (95%: 3,018-39,199) symptomatically infected adults prior to the January 23rd lockdown, respectively. Both estimates far exceed the 422 documented cases across all age groups (5). Several studies have estimated that roughly half of infections are asymptomatic (6). Thus, the number of undetected adult COVID-19 cases at the time might far exceed 10,000.

**Figure 1.**
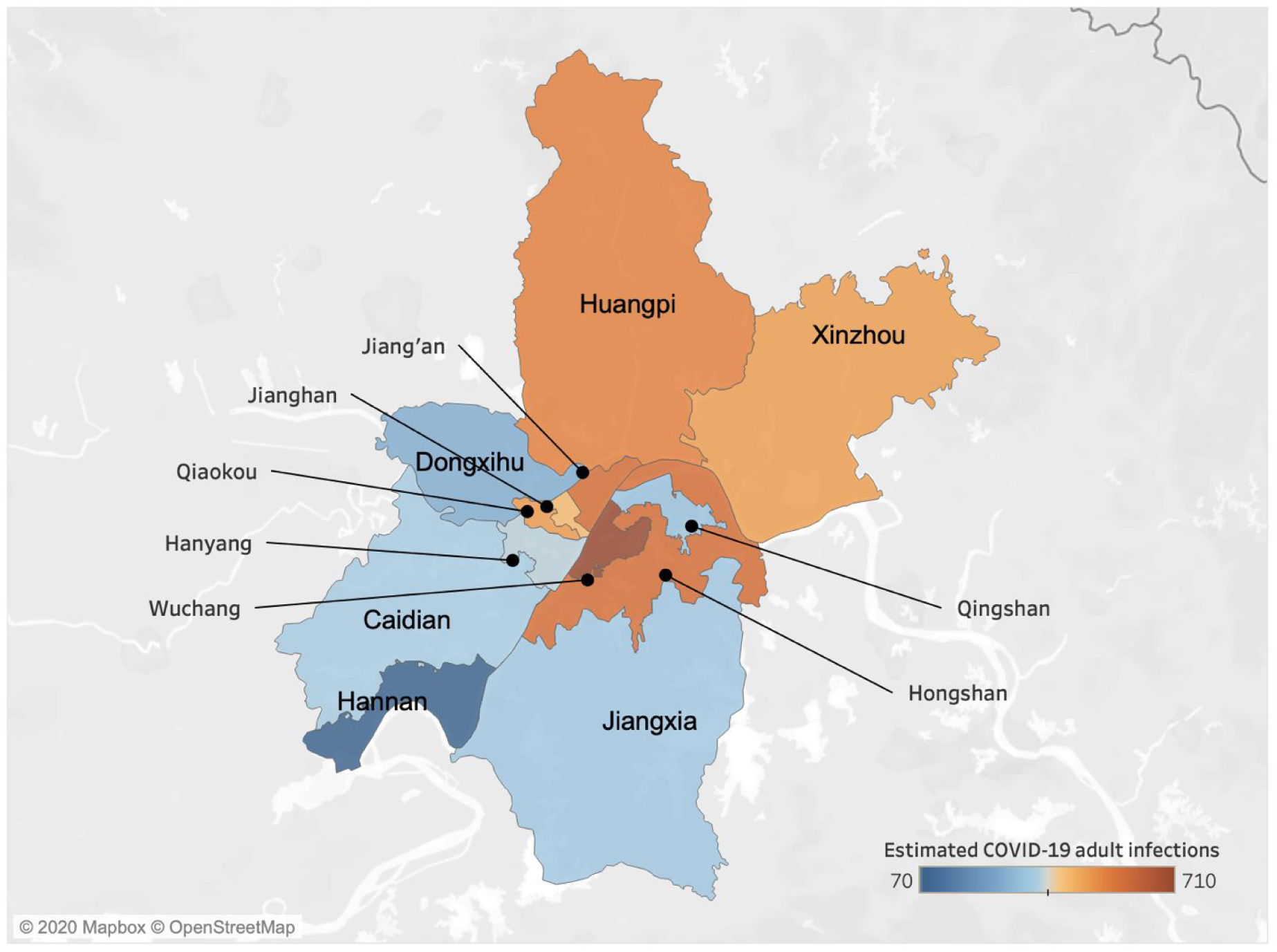
Estimated symptomatic COVID-19 infections in adults over age 30 in the 13 districts of Wuhan from December 30, 2019 to January 19, 2020. A retrospective study identified nine ILI adult cases of COVID-19 from two hospitals in central Wuhan (2). We estimate that there were a total of 5,558 [95% CI: 2,761-9,864] cases of COVID-19 in adults during that 21-day period across the 13 central districts of Wuhan, ranging from 75 cases [95% CI: 37-133] in suburban Hannan to 711 cases [95% CI: 353-1,262] in central Wuchang, as indicated by shading (Table S1).

Other data have suggested similar levels of *unseen* COVID-19 in Wuhan prior to the lockdown. For example, we previously estimated that there were 12,400 (95% CrI 3,112-58,465) total cases based on extrapolation from the timing and location of the first 19 COVID-19 cases imported from Wuhan to other countries (4). These numbers are further corroborated by a similarly-derived estimate from Imperial College of 4,000 (1,000-9,700) cases as of January 18, 2020 (7).

Using the COVID-19 to influenza test positive ratio from another retrospective study (8), we previously estimated that the number of severe but undetected *pediatric* COVID-19 cases in Wuhan prior to January 23rd, 2020 was 1105 [95% CI: 592, 1829] (9). Together, these analyses indicate that the epidemic was far more extensive than reported in January and had likely been spreading in Wuhan for several months before the lockdown. They further highlight the difficulty of determining infection fatality rates from confirmed case counts alone.

## Data Availability

Not applicable

## Acknowledgments

We acknowledge grant support from NIH (U01 GM087719).

## Author Contributions

Zhanwei Du and Lauren Ancel Meyers: conceived the study, designed statistical methods, conducted analyses, interpreted results, wrote and revised the manuscript. Benjamin J. Cowling: conceived the study, interpreted results, and revised the manuscript.

## Ethics committee approval

Not applicable.

